# Predicting Optimal Patient-Specific Postoperative Facial Landmarks for Patients with Craniomaxillofacial Deformities

**DOI:** 10.1101/2023.12.13.23299919

**Authors:** Jungwook Lee, Daeseung Kim, Xuanang Xu, Tianshu Kuang, Jaime Gateno, Pingkun Yan

## Abstract

Orthognathic surgery traditionally focuses on correcting skeletal abnormalities and malocclusion, with the expectation that an optimal facial appearance will naturally follow. However, this skeletal-driven approach can lead to undesirable facial aesthetics and residual asymmetry. To address these issues, a soft-tissue-driven planning method has been proposed. This innovative method bases bone movement estimates on the targeted ideal facial appearance, thus increasing the surgical plan’s accuracy and effectiveness. This study explores the initial phase of implementing a soft-tissue-driven approach, simulating the patient’s optimal facial look by repositioning deformed facial landmarks to an ideal state. The algorithm incorporates symmetrization and weighted optimization strategies, aligning projected optimal landmarks with standard cephalometric values for both facial symmetry and form, which are integral to facial aesthetics in orthognathic surgery. It also includes regularization to preserve the patient’s original facial characteristics. Validated using retrospective analysis of data from both preoperative patients and normal subjects, this approach effectively achieves not only facial symmetry, particularly in the lower face, but also a more natural and normalized facial form. This novel approach, aligning with soft-tissue-driven planning principles, shows promise in surpassing traditional methods, potentially leading to enhanced facial outcomes and patient satisfaction in orthognathic surgery.

## 2 Introduction

Current orthognathic surgical planning follows a skeletal-driven approach.^1–3^ It focuses on rectifying malocclusion and skeletal abnormalities, expecting optimal facial appearance to ensue. Within this framework, one can (1) trust achieving an optimal facial appearance through skeletal correction without ever simulating the soft tissue changes or (2) validate and potentially revising the skeletal plan by simulating the facial appearance using computer software. However, both tactics have limitations. On the one hand, expecting a normal facial appearance without simulating the soft-tissue deformation may overlook asymmetries within the facial soft-tissue envelope or atypical bone-to-soft-tissue relationships.^4,5^ On the other hand, simulating the facial appearance after the planned skeletal correction often necessitates time-consuming iterations and multiple plan revisions, making the process less efficient.^6–8^

To address these limitations of the current skeletal-driven method, a soft-tissue-driven planning method has been proposed.^9^ This approach estimates the necessary bone movements based on the optimal facial appearance, significantly enhancing both the efficiency and accuracy of the surgical plan.

While the accuracy in estimating an optimal facial appearance is critical for soft-tissue-driven planning, predicting this appearance before planning remains a significant challenge.^9,10^ Existing methods predominantly rely on landmark-based estimations to project postoperative facial appearance, due to the difficulties in accurately rendering the three-dimensional (3D) facial surface using limited preoperative data. These methods typically involve initial predictions of landmark movements, followed by the reconstruction of facial surfaces using simple interpolation techniques, such as thin plate spline (TPS) interpolation.^11^ Previous research has employed the partial least square (PLS) method^12^ in a supervised learning approach, using postoperative landmarks as the target. ^13–15^ These studies have incorporated data on types of deformities and surgical operations, along with preoperative and postoperative landmarks. However, the supervised approach has limitations, as it is trained to predict postoperative outcomes without guaranteeing an optimal outcome. Given that postoperative faces may still present residual deformities or asymmetries,^16^ relying solely on this data for training can be problematic. Ideally, optimal facial landmarks should adhere to universally accepted aesthetic norms represented by the distribution of cephalometric values within normal subjects while accounting for patient-specific characteristics.

The ultimate goal is to accurately estimate the optimal facial appearance for soft-tissue-driven planning, which can be achieved in two phases. The first phase involves estimating patient-specific optimal facial landmarks, and the second involves reconstructing an optimal facial surface based on these landmarks.

This study primarily focused on the first phases, addressing the significant challenge of estimating patient-specific optimal facial landmarks. The objectives were twofold: firstly, to develop an algorithm capable of accurately predicting the optimal position of facial landmarks in patients with jaw deformities; and secondly, to validate this methodology. Facial landmarks were defined as being in an optimal position when they satisfy three key outcomes: (1) perfect lower facial *symmetry*, (2) a normal facial *form*, and (3) preservation of the patient’s unique phenotype.

## 3 Materials and Methods

This study was conducted at Houston Methodist Research Institute (HMRI, Houston, Texas) and Rensselaer Polytechnic Institute (RPI, Troy, New York). The in-silico investigation utilized de-identified retrospective maxillofacial patient data. The Institutional Review Board (IRB) of HMRI approved the study—IRB# MOD00005116.

The first aim of the study was to devise an optimization algorithm to estimate the optimal facial landmarks for individuals with jaw deformities. The second aim was to validate the algorithm. To achieve both these objectives, the investigators relied on maxillofacial imaging data drawn from two distinct populations: (1) a cohort of jaw deformity patients and (2) a normal subject group.

Patients were included in the jaw deformity dataset if (1) they had undergone orthognathic surgery in the upper jaw, lower jaw, or both; (2) they had preoperative and postoperative imaging records in our virtual surgical simulation (VSP) software, AnatomicAligner (HMRI, Houston, Texas);^17^ and (3) the surgical plan had been formulated following a skeletal-driven tactic.

Subjects were included in the normal group if (1) they had no facial deformity and (2) had records in our VSP software. The VSP software files of each patient contained three-dimensional models of the facial soft-tissues, and well as their cephalometric landmarks (Table1). Infants and children were excluded from both groups.

To ensure accurate and consistent evaluation of cephalometric measurements, the 3D facial models of patients and normal subjects were aligned to their sagittal, coronal, and axial planes. The aforementioned frame of reference was calculated by the automatic function present in the AnatomicAligner software.^18^ Before the study began, the jaw deformity cohort was randomly split into two equal groups. The first group was utilized to fine-tune the optimization algorithm, while the second served to validate it.

### 3.1 Optimal Landmark Prediction

Our method for predicting optimal facial landmarks incorporates a combination of *symmetrization* process and *weighted optimization* approach. To select the appropriate measurements for the algorithm, the literature was searched to find useful published facial (i.e., soft tissue) cephalometric measurements.^19–23^ These measurements were divided into two categories: facial *symmetry* and facial *form*.

#### 3.1.1 Symmetrization

The *symmetrization* process began with the use of facial *symmetry* measurements. These measurements were subdivided into two types: *bilateral point differences* and *midpoint deviations from the sagittal plane*. The assessment of bilateral point differences involved the calculation of absolute differences in the symmetry of bilateral points across the vertical, transverse, and anteroposterior dimensions. On the other hand, the assessment of *midpoint deviations from the sagittal plane* measured the absolute perpendicular distances between jaw midline landmarks and the sagittal plane, which included the *Sn* landmark. *Symmetry* measurements are crucial for assessing the alignment and symmetry of facial features in relation to the central plane of the face.

Aiming for perfect symmetry, the *symmetrization* process adjusts bilateral points towards their average positions, effectively reducing the bilateral point differences to zero. Similarly, midpoint deviations from the sagittal plane are also aligned to zero, establishing a symmetrical baseline. In total, 12 facial *symmetry* measurements were included. (Table 2)

**Table 1.** Facial Landmarks bbreviation Full Name.

| Abbreviation | Full Name |
| --- | --- |
| *Gb'* | Soft Tissue Glabella |
| *Prn* | Pronasale |
| *CM* | Columella |
| *Sn* | Subnasale |
| *Ls* | Labiale Superius |
| *Stm* | Stomium |
| *Ch-R* | Right Cheilion |
| *Ch-L* | Left Cheilion |
| *Li* | Labiale Inferius |
| *Sl* | Sublabiale |
| *Pog'* | Soft Tissue Pogonion |
| *Me'* | Soft Tissue Menton |
| *Go'-R* | Right Soft Tissue Gonion |
| *Go'-L* | Left Soft Tissue Gonion |

**Table 2.**
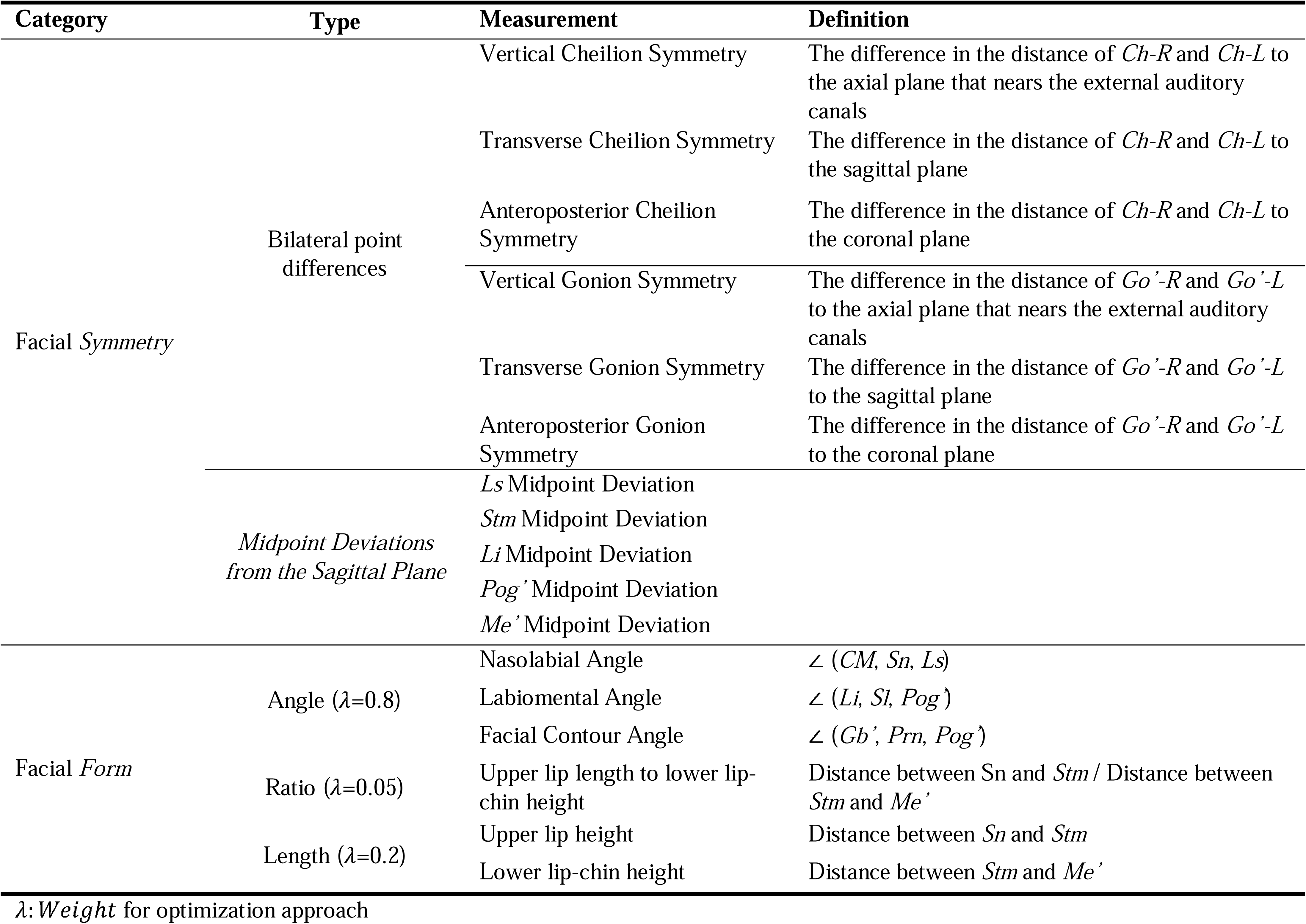
Cephalometric Measurements and Weights Used in the Optimization Approach.

#### 3.1.2 *Weighted Optimization* for Facial *Form* Measurements

To ascertain the most relevant facial *form* measurements for the algorithm, a comparative test was conducted. This analysis juxtaposed the averages and distributions of each facial *form* measurement across three distinct groups: (1) patients with jaw deformities, (2) those postoperative corrections, and (3) normal subjects. Only those *form* measurements that were altered by orthognathic surgery and subsequently aligned with the distributions of the normal group were incorporated into the optimization approach.

Facial *form* measurements were subdivided by type: *angle*, *ratio*, and *length*. This categorization was crucial because each type of measurement has its distinct units and scales. Combining them without differentiation in our model might introduce bias. To mitigate this, specific weights were allocated to each type. The weighting factor (λi) was determined through a rigorous iterative empirical process, refining the weights until the corrected cephalometric values closely matched the distribution found in the normal subject group. Data from both patient and normal subject groups were used in this determination. *Angle* measurements received a weight of 0.8, *ratio* a weight of 0.05, *length* a weight of 0.2. A total of 6 facial *form* measurements were included. (Table 2) A statistical comparison between the postoperative dataset and the normal subject dataset used for the selection is presented at Section 3.3 (statistical analysis).

The *weighted optimization* approach considered the following assumption. Given the uniqueness of each human face, its facial *form* measurements should not conform to the average of a population. Instead, optimal landmarks are those that (1) generate facial *form* measurements that are within the distribution of normal and (2) preserve the patient’s phenotype.

This approach computed the necessary displacement to rectify distorted landmarks by minimizing the objective function outlined in Equation 1.

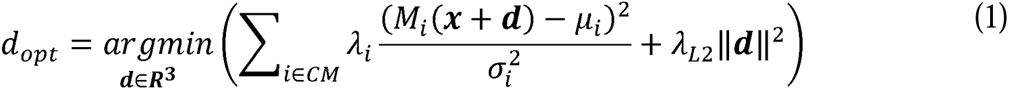

In this equation, *x* represents the deformed facial landmark, *d* is the landmark displacement vector required for the optimization, *M* is the facial *form* measurement, *CM* is a set of facial *form* measurements (Table 2), and *d_opt_* is the optimal landmark displacement. *µ* and *σ* are the mean and standard deviation of facial *form* values of the normal subject group. *λ_i_* is the weighting factor for the facial *form* type (*angle*, *ratio*, and *length*). *λ*_*L*2_ is the weighting factor for the *L*2 regularization term that preserves patient-specificity. Its value *λ*_*L*2_=1.0 was determined through an empirical process like the one used to determine *λ_i_*.

The gradient descent method was employed to minimize the objective function and find the optimal displacement vector d. The optimal facial landmarks were then predicted by applying the estimated displacement vectors d to the corresponding deformed landmarks. During the optimization, the landmarks corresponding to the upper face (Gb’, Prn, CM, and Sn) were assumed fixed because they are not directly affected by orthognathic surgery.

#### 3.1.3. Optimal Landmark Prediction

Sequential application of *symmetrization* and *weighted optimization* failed to achieve symmetry between the right and left cheilions—vertical, transverse, and anteroposterior cheilion symmetry. To solve this problem, a three-step approach was implemented. In the first step, the displacement vectors for all landmarks, excluding the right and left cheilions, were calculated by sequentially applying *Symmetrization* and *weighted optimization*. In the second step, the movement of the cheilions was inferred based on the movements of the other landmarks through Thin-Plate Spline (TPS) interpolation. The third step focused solely on the right and left cheilions, computing their displacement vectors to achieve vertical, transverse, and anteroposterior symmetry by *symmetrization*, while keeping the positions of other landmarks fixed.

### 3.2 Validation

To validate the newly proposed method, two hypotheses were formulated: (1) the new approach would yield facial landmarks that have perfect lower facial *symmetry* and normal facial *form*; (2) the new methodology would render superior results compared with outcomes obtained through established skeletal-driven planning.

A methodical procedure was employed to examine the first hypothesis, specifically whether the approach results in facial landmarks that have perfect lower facial *symmetry* and normal facial *form*. The procedure began with predicting patient-specific optimal facial landmarks for the dataset of patients exhibiting facial deformities. Subsequently, cephalometric measurements derived from these estimated facial landmarks were juxtaposed with those extracted from a dataset of normal subjects.

To scrutinize the second hypothesis, a comparative analysis was conducted between cephalometric measurements from two groups: (1) faces refined through the proposed method and (2) postoperative faces resulting from skeletal-driven planning. This comparative evaluation aimed to ascertain whether the proposed methodology offered advantages over conventional skeletal-driven planning.

In addition to testing the study hypotheses, a post-hoc test was conducted to compare the cephalometric measurements of (1) postoperative faces acquired through traditional skeletal-driven planning with those of (2) normal individuals. The purpose of this comparison was to support our assertion that skeletal-driven planning does not lead to soft-tissue normalization.

### 3.3 Statistical Analysis

For the development and validation of the new method, a rigorous statistical analysis was conducted to scrutinize the variations in the distribution of cephalometric measurements among three distinct groups: optimized preoperative, postoperative, and normal subjects. Traditional analytical approaches such as ANOVA or Kruskal-Wallis tests were deemed unsuitable for this inquiry due to the amalgamation of paired and unpaired comparisons present in the datasets. Consequently, a series of comparisons between each group was undertaken. Considering the multitude of comparisons intrinsic to this analysis, a corrected p-value of 0.017 was necessary to uphold an overall significance of 0.05. This adjustment was calculated employing the Bonferroni correction to counteract the risk of Type I error arising from multiple comparisons.

In this study, the *preoperative* and *postoperative* groups were paired, belonging to the same patients, the normal subject group was unmatched (it was a separate group of individuals). For the comparison between paired groups, each distribution was first assessed for normality. If both groups exhibited a normal distribution, a paired t-test was performed. However, if one or both groups did not follow a normal distribution, a Wilcoxon signed-rank test was used instead.

For the comparison between unpaired groups (between patient group and normal subject group), the normality of the distributions for each group was initially examined. In cases where the distributions showed normality, Levene’s test was further employed to verify the homogeneity of variances. If a significant difference in variances was detected, Welch’s t-test was applied. Otherwise, Student’s t-test was utilized. When one or both groups did not demonstrate normal distribution, the Mann-Whitney U test was employed for comparison.

## 4 Results

The deformity dataset consisted of 60 patients. Their mean age was 23.3 years, SD 6.9. Thirty-eight were females and 22 males. The normal subject group had 48 patients. Their mean age was 21.7 years, SD 2.5. Twenty-eight were females and 20 males.

As indicated in Table 3, the comparison of cephalometric measurements of the predicted optimal landmark group with those of normal subject group shows statistically significant differences for all symmetry-related measurements. Conversely, no statistically significant differences were observed for facial form measurements. This finding indicated that the predicted landmarks exhibited perfect symmetry, unlike those in the normal subject. It validated the first hypothesis, demonstrating that the proposed approach produces perfect lower face symmetry while maintaining normal facial form.

**Table 3.** Comparison of Cephalometric Measurements between Predicted Optimal Landmarks and the Normal Subjects.

| Measurement metric | Measurements from the Optimized Preoperative Group |  | Measurements from the Normal Subject Group |  | Difference |  |
| --- | --- | --- | --- | --- | --- | --- |
| | mean ± SD | 95% CI | mean ± SD | 95% CI | Test | *p*-value |
| Vertical Cheilion Sym. | 0.00±0.00 | 0.00 | 0.76±0.59 | [0, 0.87] | MWU test | <0.001* |
| Transverse Cheilion Sym. | 0.00±0.00 | 0.00 | 0.76±0.56 | [0, 0.86] | MWU test | <0.001* |
| AP Cheilion Sym. | 0.00±0.00 | 0.00 | 0.98±0.84 | [0, 1.13] | MWU test | <0.001* |
| Vertical Gonion Sym. | 0.00±0.00 | 0.00 | 2.79±1.74 | [0, 3.11] | MWU test | <0.001* |
| Transverse Gonion Sym. | 0.00±0.00 | 0.00 | 1.33±1.00 | [0, 1.52] | MWU test | <0.001* |
| AP Gonion Sym. | 0.00±0.00 | 0.00 | 2.03±1.67 | [0, 2.34] | MWU test | <0.001* |
| *Ls* Midpoint Deviation | 0.00±0.00 | 0.00 | 0.38±0.34 | [0, 0.44] | MWU test | <0.001* |
| *Stm* Midpoint Deviation | 0.00±0.00 | 0.00 | 0.49±0.38 | [0, 0.56] | MWU test | <0.001* |
| *Li* Midpoint Deviation | 0.00±0.00 | 0.00 | 0.72±0.47 | [0, 0.80] | MWU test | <0.001* |
| *Sl* Midpoint Deviation | 0.00±0.00 | 0.00 | 0.70±0.51 | [0, 0.79] | MWU test | <0.001* |
| *Pog'* Midpoint Deviation | 0.00±0.00 | 0.00 | 0.84±0.67 | [0, 0.96] | MWU test | <0.001* |
| *Me'* Midpoint Deviation | 0.00±0.00 | 0.00 | 1.25±0.97 | [0, 1.43] | MWU test | <0.001* |
| Nasolabial angle | 100.01±5.54 | [97.94, 102.07] | 101.62±9.38 | [98.90, 104.34] | Welch's t | 0.342 |
| Labiomental angle | 140.11±5.12 | [138.19, 142.02] | 137.83±10.02 | [134.92, 140.74] | Welch's t | 0.190 |
| Facial contour angle | 167.66±3.34 | [166.42, 168.91] | 166.86±4.43 | [144.6, 146.8] | Ind t | 0.395 |
| Upper lip length to lower lip-chin height ratio | 0.46±0.04 | [0.45, 0.48] | 0.48±0.05 | [0.46, 0.50] | Welch's t | 0.120 |
| Upper lip length | 21.44±1.35 | [20.93, 21.94] | 22.07±2.08 | [21.47, 22.67] | Welch's t | 0.106 |
| Lower lip-chin height | 46.30±2.97 | [45.19, 47.40] | 46.17±3.67 | [45.11, 47.24] | Ind t | 0.877 |

Sym: symmetry, AP: anteroposterior, SD: standard deviation, CI: confidence interval, MWU test: Mann-Whitney U test, Welch's t: Welch's t-test, Ind t: Independent t-test, \* Significant difference ( $p<0.017$ ).

Similarly, when comparing the cephalometric measurements obtained from the predicted optimal landmarkgroup with those of postoperative patients (as presented in Table 4), it was revealed that all symmetry measurements exhibited statistically superior results in the predicted optimal landmark. Additionally, the facial form measurements revealed no significant differences among the predicted optimal landmark group, postoperative group, and normal subject group.

**Table 4.** Comparison of Cephalometric Measurements between Predicted Optimal Landmarks and Postoperative Patients.

| Measurement Metric | Measurements from the<br>Optimized Preoperative Group |  | Measurements from the<br>Postoperative Group |  | Difference |  |
| --- | --- | --- | --- | --- | --- | --- |
| | mean ± SD | 95% CI | mean ± SD | 95% CI | Test | *p*-value |
| Vertical Cheilion Sym. | 0.00±0.00 | 0.00 | 1.13±0.94 | [0, 1.34] | WSR test | <0.001* |
| Transverse Cheilion Sym. | 0.00±0.00 | 0.00 | 0.95±0.93 | [0, 1.17] | WSR test | <0.001* |
| AP Cheilion Sym. | 0.00±0.00 | 0.00 | 1.20±0.81 | [0, 1.38] | WSR test | <0.001* |
| Vertical Gonion Sym. | 0.00±0.00 | 0.00 | 5.04±3.41 | [0, 5.82] | WSR test | <0.001* |
| Transverse Gonion Sym. | 0.00±0.00 | 0.00 | 1.51±1.17 | [0, 1.78] | WSR test | <0.001* |
| AP Gonion Sym. | 0.00±0.00 | 0.00 | 3.97±3.25 | [0, 4.72] | WSR test | <0.001* |
| *Ls* Midpoint Deviation | 0.00±0.00 | 0.00 | 0.55±0.50 | [0, 0.66] | WSR test | <0.001* |
| *Stm* Midpoint Deviation | 0.00±0.00 | 0.00 | 0.75±0.65 | [0, 0.90] | WSR test | <0.001* |
| *Li* Midpoint Deviation | 0.00±0.00 | 0.00 | 1.12±0.96 | [0, 1.34] | WSR test | <0.001* |
| *Sl* Midpoint Deviation | 0.00±0.00 | 0.00 | 1.46±1.19 | [0, 1.74] | WSR test | <0.001* |
| *Pog'* Midpoint Deviation | 0.00±0.00 | 0.00 | 2.08±1.66 | [0, 2.46] | WSR test | <0.001* |
| *Me'* Midpoint Deviation | 0.00±0.00 | 0.00 | 2.53±1.93 | [0, 2.97] | WSR test | <0.001* |
| Nasolabial angle | 100.01±5.54 | [97.94, 102.07] | 102.34±9.29 | [98.87, 105.80] | Paired t | 0.024 |
| Labiomental angle | 140.11±5.12 | [138.19, 142.02] | 135.88±10.40 | [132.00, 139.76] | Paired t | 0.027 |
| Facial contour angle | 167.66±3.34 | [166.42, 168.91] | 169.03±6.57 | [166.58, 171.49] | Paired t | 0.117 |
| Upper lip length to lower lip-chin height ratio | 0.46±0.04 | [0.45, 0.48] | 0.47±0.06 | [0.45, 0.50] | Paired t | 0.169 |
| Upper lip length | 21.44±1.35 | [20.93, 21.94] | 21.69±2.26 | [20.85, 22.53] | Paired t | 0.454 |
| Lower lip-chin height | 46.30±2.97 | [45.19, 47.40] | 45.74±3.39 | [44.48, 47.01] | Paired t | 0.117 |

Sym: symmetry AP: anteroposterior, SD: standard deviation, CI: confidence interval, WSR test: Wilcoxon Signed Rank test, Paired t: Paired t-test, \* Significant difference ( $p < 0.017$ ).

**Table 5.** Comparison of Cephalometric Measurements between Postoperative Patients and Normal Subjects.

| Measurement metric | Measurements from the Postoperative Group |  | Measurements from the Normal Subject Group |  | Difference |  |
| --- | --- | --- | --- | --- | --- | --- |
| | mean ± SD | 95% CI | mean ± SD | 95% CI | Test | *p*-value |
| Vertical Cheilion Sym. | 1.13±0.94 | [0, 1.34] | 0.76±0.59 | [0, 0.87] | MWU test | 0.130 |
| Transverse Cheilion Sym. | 0.95±0.93 | [0, 1.17] | 0.76±0.56 | [0, 0.86] | Ind t | 0.307 |
| AP Cheilion Sym. | 1.20±0.81 | [0, 1.38] | 0.98±0.84 | [0, 1.13] | MWU test | 0.174 |
| Vertical Gonion Sym. | 5.04±3.41 | [0, 5.82] | 2.79±1.74 | [0, 3.11] | Welch's t | 0.002* |
| Transverse Gonion Sym. | 1.51±1.17 | [0, 1.78] | 1.33±1.00 | [0, 1.52] | MWU test | 0.548 |
| AP Gonion Sym. | 3.97±3.25 | [0, 4.72] | 2.03±1.67 | [0, 2.34] | MWU test | 0.001* |
| *Ls* Midpoint Deviation | 0.55±0.50 | [0, 0.66] | 0.38±0.34 | [0, 0.44] | MWU test | 0.201 |
| *Stm* Midpoint Deviation | 0.75±0.65 | [0, 0.90] | 0.49±0.38 | [0, 0.56] | MWU test | 0.108 |
| *Li* Midpoint Deviation | 1.12±0.96 | [0, 1.34] | 0.72±0.47 | [0, 0.80] | Welch's t | 0.040 |
| *Sl* Midpoint Deviation | 1.46±1.19 | [0, 1.74] | 0.70±0.51 | [0, 0.79] | MWU test | 0.004* |
| *Pog'* Midpoint Deviation | 2.08±1.66 | [0, 2.46] | 0.84±0.67 | [0, 0.96] | MWU test | <0.001* |
| *Me'* Midpoint Deviation | 2.53±1.93 | [0, 2.97] | 1.25±0.97 | [0, 1.43] | MWU test | <0.001* |
| Nasolabial angle | 102.34±9.29 | [98.87, 105.80] | 101.62±9.38 | [98.90, 104.34] | Ind t | 0.743 |
| Labiomental angle | 135.88±10.40 | [132.00, 139.76] | 137.83±10.02 | [134.92, 140.74] | Ind t | 0.412 |
| Facial contour angle | 169.03±6.57 | [166.58, 171.49] | 166.86±4.43 | [144.6, 146.8] | Welch's t | 0.116 |
| Upper lip length to lower lip-chin height ratio | 0.47±0.06 | [0.45, 0.50] | 0.48±0.05 | [0.46, 0.50] | Ind t | 0.779 |
| Upper lip length | 21.69±2.26 | [20.85, 22.53] | 22.07±2.08 | [21.47, 22.67] | Ind t | 0.449 |
| Lower lip-chin height | 45.74±3.39 | [44.48, 47.01] | 46.17±3.67 | [45.11, 47.24] | Ind t | 0.607 |

Sym: symmetry, AP : anteroposterior, SD: standard deviation, CI:confidence interval, MWU test: Mann-Whitney U test, Welch's t: Welch's t-test, Ind t: Independent t-test, \* Significant difference ( $p<0.017$ ).

This outcome provides confirmation for our second hypothesis, proving that the new methodology yields superior results compared to outcomes achieved through established bone-driven planning, especially in the context of enhancing facial symmetry without the degradation of facial form.

The post-hoc comparison between the cephalometric measurements of postoperative patients and those of normal subjects showed that that 5 out of 14 measurements were statistically worse in the postoperative group. Again, all statistically significant differences in cephalometric measurements pertain to facial symmetry. This outcome confirms our assertion that bone-driven planning does not lead to complete soft-tissue normalization, particularly in facial symmetry.

Figures 1 and 2 provide visual representations of these findings. Figure 1 displays the distribution of each measurement across all groups, including cephalometric measurements of the preoperative group to illustrate changes following surgery. Figure 2 presents an example case demonstrating the estimated optimal landmarks.

**Figure 1.**
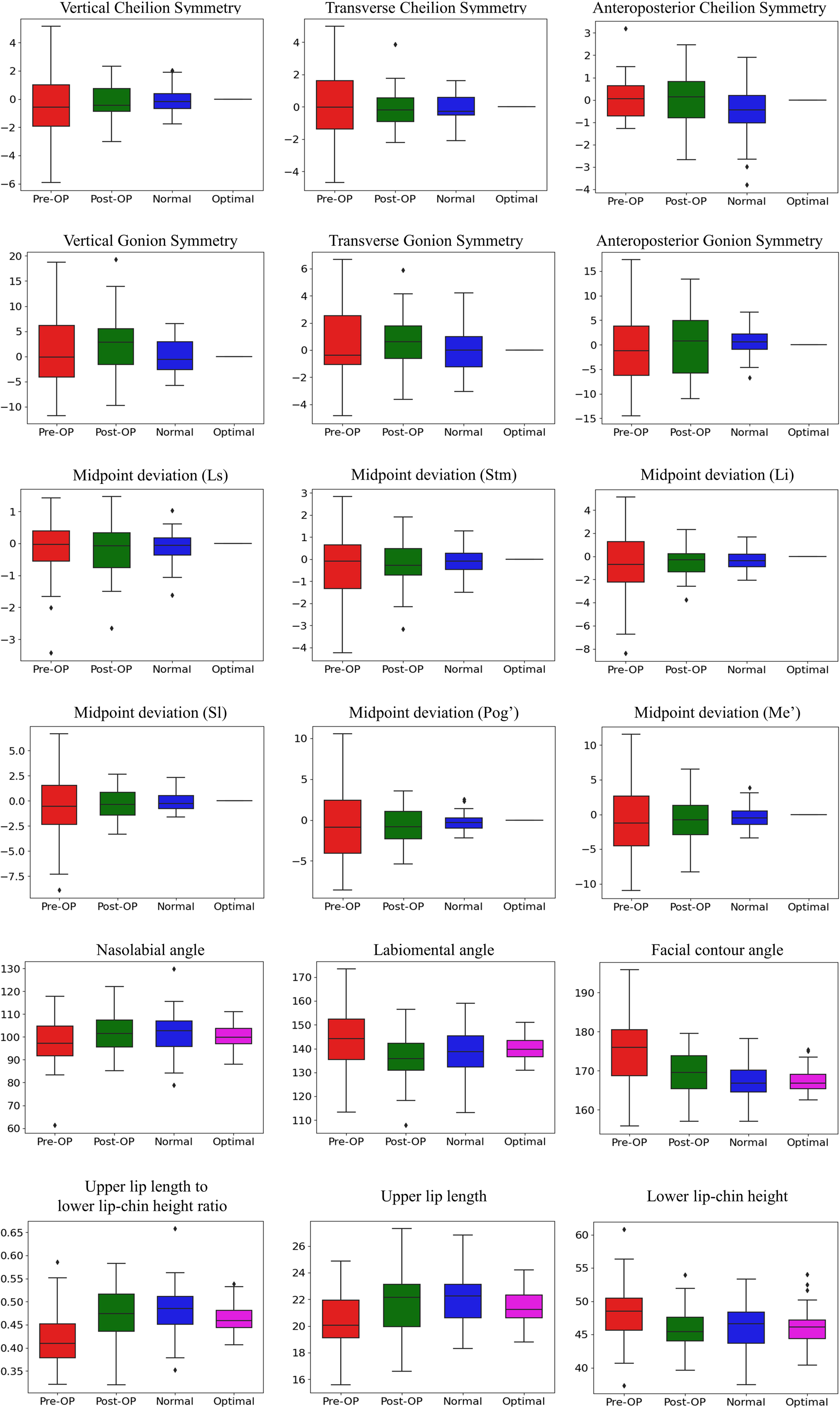
Box plots of cephalometric measurement of preoperative (Pre-OP; red), postoperative (Post-OP; green), normal group (blue), and predicted optimal landmarks (magenta).

**Figure 2.**
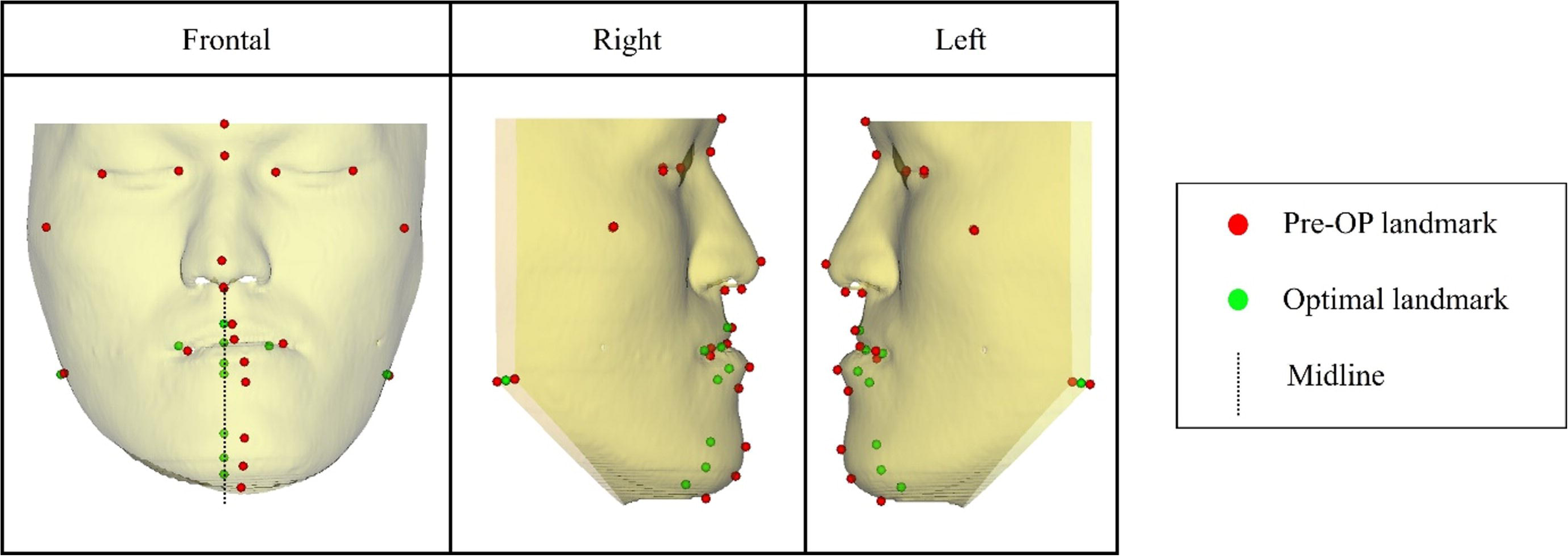
The positions of pre-operative (Pre-OP) and transformed landmarks in the frontal view (left), right profile view (middle), and left profile view (right) of randomly selected patient. Midline is defined as vertical line passing through the *subnasale*.

## 5 Discussion

In this study, the researchers have pioneered an innovative approach for the prediction of optimal facial landmarks in individuals suffering from jaw deformities. The **key findings** of the study are significant in several aspects. Firstly, the developed methodology achieves facial symmetry, particularly in the lower face, while ensuring that the facial appearance remains patient-specific and normal. Secondly, this novel approach potentially surpasses traditional bone-driven planning methods in delivering enhanced facial outcomes, with notable improvement in symmetry.

Furthermore, a post-hoc analysis comparing cephalometric measurements of *postoperative patients* to those of *normal subjects* highlighted that traditional bone-centric planning often falls short in achieving complete soft-tissue normalization, especially in terms of facial *symmetry*.

The **clinical relevance** of this project lies in its challenge to the current skeletal-centric paradigm in orthognathic surgery. The prevalent skeletal-centric approach emphasized the correction of malocclusion and skeletal anomalies, with the expectation that aesthetically pleasing facial appearance would ensue. During planning, skeletal-centric method is implemented in two distinct ways. Firstly, some clinicians assume that correcting skeletal deformities alone will result in optimal facial aesthetics, hence they do not simulate the soft tissue changes. Secondly, other clinicians employ computer algorithms to predict the facial outcome post-skeletally corrective procedures, thus substantiating the skeletal plan.

Despite the everyday use of these methodologies, practical limitations are evident. Relying solely on skeletal adjustments without visualizing the soft tissue changes could miss asymmetries in the facial soft tissue or atypical correlations between bone structure and adjacent soft tissues.^4,5^ Conversely, the employment of facial simulation software in the planning stage, while beneficial for visualizing postoperative outcomes, often necessitated labor-intensive iterative processes and multiple revisions of the surgical plan. This dichotomy highlights the inherent complexities and challenges in achieving a harmonious balance between skeletal correction and desirable facial aesthetics in orthognathic surgery.

The research group proposed a novel soft-tissue-driven planning method for orthognathic surgery,^9^ which could potentially resolve the aforementioned issues. This method began by simulating an optimal facial appearance and then calculated the necessary skeletal framework to support the overlying soft tissues, considering their thickness and composition. The process culminates in guiding the three-dimensional alignment of the jaw segments to match the ideal skeletal framework closely. This step goes beyond mere aesthetic alignment; it ensures the maintenance of normal occlusion and jaw function for the patient. By emphasizing the role of soft tissue in surgical planning, the soft-tissue-driven method aims to achieve outcomes that are not only aesthetically pleasing but also functionally sound. This represents a shift from traditional methods that might focus primarily on the skeletal structure, offering a more patient-centric and comprehensive approach to orthognathic surgery.

This study tackles the first stage necessary for applying a soft-tissue-driven approach, aiming to simulate the optimal facial appearance of patients. This task involves two key steps: firstly, repositioning deformed facial landmarks to an ideal position, and secondly, rendering the optimal soft-tissue surface. In this paper, we propose a solution for the initial step, with plans to address the second step in a subsequent study.

Rather than relocating facial landmarks to positions typical of an average population, our weighted optimization approach moves deformed facial landmarks to appropriate positions while preserving each patient’s unique characteristics. Our method differentiates between cephalometric measurements for evaluating symmetry and those for assessing facial *form*. The method aims at perfect lower facial *symmetry* but avoids average facial *form*. Although achieving perfect lower facial *symmetry* is not surgically feasible, creating a symmetrical template for planning is valuable. It may decrease the likelihood of postoperative asymmetry.

On the other hand, an average facial *form* might not be suitable for all patients. Typically, cephalometric measurements in a normal population are distributed around a mean value. By limiting the movement of facial landmarks to positions that enter the normal range, but are not necessarily aligned with the means, one can maintain the patient’s phenotype and enhance the likelihood that the surgery will be feasible.

Despite the promising results, our study has **limitations**. Firstly, the accuracy and generalizability of the approach depends on the characteristics of the normal group population. To enhance the applicability of the method, future research will focus on expanding the normative database to include a more diverse population, considering factors such as gender, age, and ethnicity.

Another limitation lies in the heavy reliance on cephalometric measurements as the primary measure for the optimal landmark prediction. While cephalometric measurements provide valuable information for assessing facial aesthetics, they have inherent limitations in capturing the complex multidimensional nature of facial aesthetics. To overcome this limitation, future studies could explore the integration of additional measurements utilizing three-dimensional information.

Finally, while in the study, the distributions of cephalometric measurements in the *estimated preoperative group* are within those of the *postoperative group* (as shown in Figure 1), the study does not prove that the *predicted optimal facial appearances* are surgically attainable for individual patients.

The **future direction** of this project involves developing all the necessary technology for implementing soft-tissue-driven planning. This includes: (1) rendering an optimal facial appearance based on optimized landmarks, (2) calculating the necessary skeletal framework to support the overlying soft tissues, and (3) guiding the three-dimensional alignment of the jaw segments to closely match the idealized skeletal framework.

In **conclusion**, the novel approach for predicting optimal facial landmarks achieved an optimal balance between normalization of facial deformities and preservation of individual characteristics. The new method signifies a substantial advancement in optimal face prediction for soft-tissue-driven surgical planning, holding the promise of enhancing surgical outcomes and patient satisfaction.

## Declarations

### Ethical approval

The study was approved by the institutional review boards of Houston Methodist Hospital and Research Institute (IRB#: MOD00005116)

### Patient consent

Not applicable.

## Data Availability

The data that support the findings of this study are not publicly available due to ethical restrictions imposed by the MOD00005116.

## 6 References

1. Seo HJ., Choi Y-K. Current trends in orthognathic surgery. Arch Craniofac Surg 2021;22(6):287.

2. Choi J-W., Lee JY., Choi J-W., Lee JY. Treatment Strategy for Class II Orthognathic Surgery: Orthodontic Perspective. The Surgery-First Orthognathic Approach: With Discussion of Occlusal Plane-Altering Orthognathic Surgery 2021:71–100.

3. Hammoudeh JA., Howell LK., Boutros S., Scott MA., Urata MM. Current status of surgical planning for orthognathic surgery: traditional methods versus 3D surgical planning. Plast Reconstr Surg Glob Open 2015;3(2).

4. Kim D., Kuang T., Rodrigues YL., Gateno J., Shen SGF., Wang X., et al. A new approach of predicting facial changes following orthognathic surgery using realistic lip sliding effect. Medical Image Computing and Computer Assisted Intervention–MICCAI 2019: 22nd International Conference, Shenzhen, China, October 13–17, 2019, Proceedings, Part V 22. Springer; 2019. p. 336–44.

5. Bell WH., Ferraro JW. Modern practice in orthognathic and reconstructive surgery. Plast Reconstr Surg 1993;92(2):362.

6. Ma L., Xiao D., Kim D., Lian C., Kuang T., Liu Q., et al. Simulation of postoperative facial appearances via geometric deep learning for efficient orthognathic surgical planning. IEEE Trans Med Imaging 2022;42(2):336–45.

7. Schendel SA., Jacobson R., Khalessi S. 3-dimensional facial simulation in orthognathic surgery: is it accurate? Journal of Oral and Maxillofacial Surgery 2013;71(8):1406–14.

8. Rokhshad R., Keyhan SO., Yousefi P. Artificial intelligence applications and ethical challenges in oral and maxillo-facial cosmetic surgery: a narrative review. Maxillofac Plast Reconstr Surg 2023;45(1):14.

9. Fang X., Kim D., Xu X., Kuang T., Lampen N., Lee J., et al. Soft-Tissue Driven Craniomaxillofacial Surgical Planning. International Conference on Medical Image Computing and Computer-Assisted Intervention. Springer; 2023. p. 186–95.

10. Modabber A., Baron T., Peters F., Kniha K., Danesh G., Hölzle F., et al. Comparison of soft tissue simulations between two planning software programs for orthognathic surgery. Sci Rep 2022;12(1):5013.

11. Keller W., Borkowski A. Thin plate spline interpolation. J Geod 2019;93:1251–69.

12. Wold H. Estimation of principal components and related models by iterative least squares. Multivariate Analysis 1966:391–420.

13. Suh H-Y., Lee S-J., Lee Y-S., Donatelli RE., Wheeler TT., Kim S-H., et al. A more accurate method of predicting soft tissue changes after mandibular setback surgery. Journal of Oral and Maxillofacial Surgery 2012;70(10):e553–62.

14. Yoon K-S., Lee H-J., Lee S-J., Donatelli RE. Testing a better method of predicting postsurgery soft tissue response in Class II patients: a prospective study and validity assessment. Angle Orthod 2015;85(4):597– 603.

15. Suh H-Y., Lee H-J., Lee Y-S., Eo S-H., Donatelli RE., Lee S-J. Predicting soft tissue changes after orthognathic surgery: the sparse partial least squares method. Angle Orthod 2019;89(6):910–6.

16. Ajmera DH., Hsung RT-C., Singh P., Wong NSM., Yeung AWK., Lam WYH., et al. Three-dimensional assessment of facial asymmetry in Class III subjects. Part 1: a retrospective study evaluating postsurgical outcomes. Clin Oral Investig 2022;26(7):4947–66.

17. Yuan P., Mai H., Li J., Ho DC-Y., Lai Y., Liu S., et al. Design, development and clinical validation of computer-aided surgical simulation system for streamlined orthognathic surgical planning. Int J Comput Assist Radiol Surg 2017;12:2129–43.

18. Gateno J., Jajoo A., Nicol M., Xia JJ. The primal sagittal plane of the head: a new concept. Int J Oral Maxillofac Surg 2016;45(3):399–405.

19. Naini FB., Gill DS. Facial aesthetics: 2. Clinical assessment. Dent Update 2008;35(3):159–70.

20. Jagadish Chandra H., Ravi MS., Sharma SM., Rajendra Prasad B. Standards of facial esthetics: an anthropometric study. J Maxillofac Oral Surg 2012;11:384–9.

21. Oh J., Han JJ., Ryu S-Y., Oh H-K., Kook M-S., Jung S., et al. Clinical and cephalometric analysis of facial soft tissue. Journal of Craniofacial Surgery 2017;28(5):e431–8.

22. Eggerstedt M., Rhee J., Urban MJ., Mangahas A., Smith RM., Revenaugh PC. Beauty is in the eye of the follower: facial aesthetics in the age of social media. Am J Otolaryngol 2020;41(6):102643.

23. Reyneke JP., Ferretti C. Diagnosis and planning in orthognathic surgery. Oral and Maxillofacial Surgery for the Clinician 2021:1437–62.

